# Overexpression of Fibroblast Activation Protein (FAP) in stroma of proliferative inflammatory atrophy (PIA) and primary adenocarcinoma of the prostate

**DOI:** 10.1101/2024.04.04.24305338

**Authors:** Fernanda Caramella-Pereira, Qizhi Zheng, Jessica L. Hicks, Sujayita Roy, Tracy Jones, Martin Pomper, Lizamma Antony, Alan K. Meeker, Srinivasan Yegnasubramanian, Angelo M. De Marzo, W. Nathaniel Brennen

**Author notes:** indicate co-senior authors (Angelo M. De Marzo, W. Nathaniel Brennen). Address for correspondence: A M De Marzo, MD, Department of Pathology, 1550 Orleans Street. CRB2, Rm144, Baltimore, Maryland, Main.

## Abstract

Fibroblast activation protein (FAP) is a serine protease upregulated at sites of tissue remodeling and cancer that represents a promising therapeutic and molecular imaging target. In prostate cancer, studies of FAP expression using tissue microarrays are conflicting, such that its clinical potential is unclear. Furthermore, little is known regarding FAP expression in benign prostatic tissues. Here we demonstrated, using a novel iterative multiplex IHC assay in standard tissue sections, that FAP was nearly absent in normal regions, but was increased consistently in regions of proliferative inflammatory atrophy (PIA). In carcinoma, FAP was expressed in all cases, but was highly heterogeneous. High FAP levels were associated with increased pathological stage and cribriform morphology. We verified that FAP levels in cancer correlated with CD163+ M2 macrophage density. In this first report to quantify FAP protein in benign prostate and primary tumors, using standard large tissue sections, we clarify that FAP is present in all primary prostatic carcinomas, supporting its potential clinical relevance. The finding of high levels of FAP within PIA supports the injury/regeneration model for its pathogenesis and suggests that it harbors a protumorigenic stroma. Yet, high levels of FAP in benign regions could lead to false positive FAP-based molecular imaging results in clinically localized prostate cancer.

## Introduction

Fibroblast activation protein (FAP) is a type II transmembrane serine protease with both endopeptidase and dipeptidyl peptidase (DPP) activities.^1–3^ FAP expression is generally absent in normal tissues but is upregulated at sites of tissue remodeling during wound healing, in regions of fibrosis and in many invasive cancers. ^1,3–5^ FAP has been implicated in cancer initiation and progression by promoting tumor cell proliferation, angiogenesis, stromal invasion and immune suppression. ^1,3,6^ As a result, FAP protease activity has emerged as a molecular imaging target in cancer ^7–9^ and as a potential anti-cancer drug target.^1,10^ In addition, FAP activity is a promising target for prodrugs designed to be activated in regions of high FAP expression. ^11,12^

In prostate cancer there are conflicting studies regarding FAP expression.^9,13–16^ Using multiplex immunofluorescence on tissue microarrays (TMAs), increasing FAP levels have been positively associated with higher Gleason score, MRI positivity, biochemical recurrence, and disease progression after prostatectomy.^16^ However, a separate recent study found no FAP protein expression in primary tumors using TMAs, but noted high levels of expression in metastatic castration resistant prostate cancer (mCRPC).^13^ Furthermore, while FAP mRNA has been examined in relation to cribriform morphology, ^15,17^ FAP protein expression has not been systematically characterized in relation to morphological features of disease aggressiveness beyond Gleason grading in primary prostate carcinoma. Finally, while FAP expression was found to be increased in a region of inflammation in benign prostatic hyperplasia (BPH) in a single case, ^9^ its expression in benign prostatic tissues has been reported as negative or has not been commented upon and it has not been systematically studied in benign inflammatory lesions of the prostate. ^9,13–16^

The lack of information regarding FAP expression in benign prostate tissues represents an important knowledge gap regarding both prostatic carcinogenesis and FAP imaging and theranostics. For example, non-cancerous regions in the prostate contain foci of inflammation, epithelial cell injury, and proliferation, referred to as proliferative inflammatory atrophy (PIA). To date, FAP expression has not been examined in these lesions. These foci have been proposed as precursor lesions for the development of high grade prostatic intraepithelial neoplasia (PIN) and/or invasive adenocarcinoma.^18–20^ Also, most studies of PIA have focused on the epithelial cells. For example, in addition to an increased proliferative fraction, intermediate luminal cells in PIA show increased expression of GSTP1 and BCL-2, ^21^ GSTA1, ^22^ Keratin 5, ^23^ PTGS2, ^24,25^ and lactoferrin (LTF). ^26^ Luminal intermediate cells in PIA also show decreased expression of CD38,^27^ rare *GSTP1* methylation, ^28,29^ rare *TMPRSS2-ERG* gene fusions, ^30^ as well as increased expression of genes indicative of immune activation, some of which have overlap with those seen in lung club cells. ^31,32^ These findings provide strong evidence that many luminal epithelial cells in PIA are experiencing a stress response, and/or immune activation response, yet no previous studies have focused on the stromal compartment in PIA where FAP would be expected to be expressed. It is also important to evaluate the expression pattern and levels of FAP in non-neoplastic prostatic tissues, since the presence of high levels of FAP in such benign regions could lead to false positive FAP-based molecular imaging results. To address these unresolved questions in a spatially relevant and quantitative manner, we developed a novel quantitative chromogenic multiplex IHC assay centered around a highly analytically validated FAP antibody for IHC. Since FAP expression tends to be heterogeneous, we used whole slide tissue sections from radical prostatectomy specimens to avoid under sampling of tissue that may occur using TMAs.

## Materials and Methods

### Patient Samples

Formalin fixed and paraffin embedded (FFPE) prostate tissue sections were obtained from The Johns Hopkins Hospital department of pathology from radical prostatectomy specimens from men treated for clinically localized prostatic adenocarcinoma. The use of these tissues was approved by the Johns Hopkins University School of Medicine Institutional Review Board (NA_00087094). Prostatectomy specimens were handled as previously described. ^33^ For each case (N=56), a single FFPE block was selected representing the highest grade tumor. The Gleason grade groups of the retrieved cases ranged from GG1 to GG5, and the pathological stages ranged from organ confined (T2,N0) to pelvic lymph node metastasis (any T, N1) (**Table 1**).

**Table 1.** Grade Group and Stage at Prostatectomy.

| Grade Group | T2N0 | T3N0 | T3BN0/N1 | Total |
| --- | --- | --- | --- | --- |
| 1 | 6 | 0 | 0 | 6 |
| 2 | 13 | 8 | 1 | 22 |
| 3 | 4 | 4 | 2 | 10 |
| 4 | 3 | 0 | 0 | 3 |
| 5 | 2 | 9 | 4 | 15 |
| Total | 28 | 21 | 7 | 56 |

### IHC

Antibodies used in this study included those against FAP, CD45, CD163, p63, Keratin 8 (CK8), desmin, CD31, vimentin, s100 and alpha smooth muscle actin; the details and conditions are presented in **Supplemental Table 1**. Each antibody was optimized by individual single-plex staining before being incorporated into the iterative multiplex IHC assay as described.^33^ These antibodies were combined in two different panels using a multiplex approach that entails chromogenic IHC staining followed by whole slide scanning, chromogen and antibody stripping, and then iterating with the next antibody and cycle.

Slides were scanned using a 40x objective on a Roche/Ventana DP200. Whole slide images (WSIs) from each staining round were uploaded to the HALO image analysis platform (Indica Labs), and processed by color deconvolution, image registration and image fusion as described.^33^ CK8 staining was used to facilitate training of a random forest classifier to determine epithelial and stromal areas in regions of tumor as described.^33^ HALO Area Quantification (Area Quantification FL v2.3.4) tools were used to determine areas of positive staining for FAP, CD163 and for CD45.

### Quantification

Regions of normal appearing epithelium and stroma, PIA and invasive adenocarcinoma were annotated on whole slide images of H&E stained sections. For quantifying FAP, we used two separate threshold cutoffs for positive pixels. The lower cutoff was used to identify all positive staining above background and the higher cutoff was used to only quantify high intensity areas of FAP. For each of these thresholds, we defined the percent area of FAP staining (FAP Area Fraction) as the percentage of the FAP positive pixel counts divided by the total pixel counts in the annotated total area. The total tumor area, consisting of both tumor epithelium and tumor stroma, was defined by manual annotation of tumor regions, excluding benign glands and after subtracting lumens and empty spaces. The percent stromal area of FAP straining (FAP Stroma Area Fraction) was defined as the percentage of area of FAP positive pixel counts divided by the total stromal area. In general, both of these FAP threshold measurements provided similar trends overall and the results correlated with each other.

For quantification of M2 macrophages, we used the positive pixel counts of the CD163 area to generate a CD163 Area Fraction and CD163 Stroma Area Fraction for each case as above. For total leukocytes, we used the positive pixel counts of the CD45 area to generate a CD45 Area Fraction and CD45 Stroma Area Fraction.

#### In Situ Hybridization for FAP

RNA in situ hybridization (RISH) was performed using the RNAscope 2.5 FFPE Brown Reagent Kit (Advanced Cell Diagnostics, Inc.) largely as previously described.^34^ Briefly, FFPE cell line slides were baked at 60°C for 30 minutes followed by deparaffinization in 100% xylene twice for 5 minutes each and two changes of 100% alcohol. The slides were treated with endogenous peroxidase-blocking pretreatment reagent and then incubated for 18 minutes in a boiling water bath and then treated with protease digestion buffer for 20 minutes at 40°C. The slides were incubated with ACD RNAscope target probe set Cat No. 411971, NCBI reference accession No. NM_004460.2, target region (Base pairs: 486-1588) for 2 hours at 40°C, followed by signal amplification. DAB was used for colorimetric detection for 10 minutes at room temperature.

### hPrCSC-44c immortalization

PrCSC-44c were obtained from radical prostatectomy tissue as previously described.^35^ PrCSC-44c was hTERT-immortalized using the pLOX-TERT-iresTK (plasmid #12245, Addgene, Watertown MA) lentiviral vector^36^ as previously described to generate hPrCSC-44c as a model of prostate cancer-derived stromal cells (MSCs + CAFs).^37^ Briefly, HEK293T cells were first co-transfected with the TERT-containing vector and packaging plasmid using Lipofectamine 2000 Transfection reagent (Sigma-Aldrich, St. Louis, MO) diluted in Opti-MEM I reduced serum media (Gibco, Waltham, MA). After 24 hours at 37°C, the media was removed and replaced with standard Opti-MEM. The supernatant was collected after an additional 24 hours of incubation at 37°C, and the virus was harvested and concentrated by ultracentrifugation for 2 hours at 100,000 x g. Virus was then added to the PrCSC-44c culture media (hMSC Expansion media, RoosterBio, Frederick, MD) supplemented with 8 mg/mL of polybrene (Sigma-Aldrich) and incubated at 37°C for 24 hours. The virus-containing supernatant was removed and replaced with fresh media. Immortalization was confirmed via continuous passaging of the cells at ∼80% confluence in hMSC Expansion media at a 1:5 dilution and the population doublings significantly exceeded those of the non-immortalized parental cells as they underwent senescence under standard tissue culture conditions (18 vs. >30 passages).

### Statistical analysis

Statistical analysis was carried out in STATA 18 for Microsoft Windows. Differences in percent FAP area between groups were assessed using the Wilcoxon rank-sum (Mann-Whitney) test for unpaired analyses and the Wilcoxon signed-rank test for paired analyses.

Statistical significance was set at P<0.05. Linear regression was used to examine the relationship between the percent FAP area and percent CD163 area and percent CD45 areas.

## Results

### Analytic Validation of FAP IHC Assay and Multiplex Staining

To analytically validate an IHC assay for FAP, cell lines with known FAP expression were processed by formalin fixation and paraffin embedding (FFPE)^38^ and stained with an anti-FAP antibody. As a positive control, we used a cell line derived from TERT-immortalized prostate stromal cells (hPrCSC-44c cells) that expresses FAP mRNA and protein, ^12,35^ and these cells were positive by IHC (**Fig. 1A**). As an isogenic negative control, hPrCSC-44c cells were subjected to targeted disruption of *FAP* by CRISPR/Cas9. Staining for FAP in these cells was negative (**Fig. 1B**). EnzaR-CWR22Rv1^39,40^ prostate cancer cells do not express FAP mRNA and protein,^12^ and they were negative by IHC (**Fig. 1C**) and by *in situ* hybridization for FAP mRNA (**Fig. 1G**). As an additional isogenic control, we transfected a cDNA clone encoding human FAP into EnzaR-CWR22RV1 cells and this resulted in robust anti-FAP immunostaining (**Fig. 1D**) and mRNA expression (**Fig 1H**). Taken together, these results provide genetic validation of our IHC assay conditions for FAP protein.

**Figure 1.**
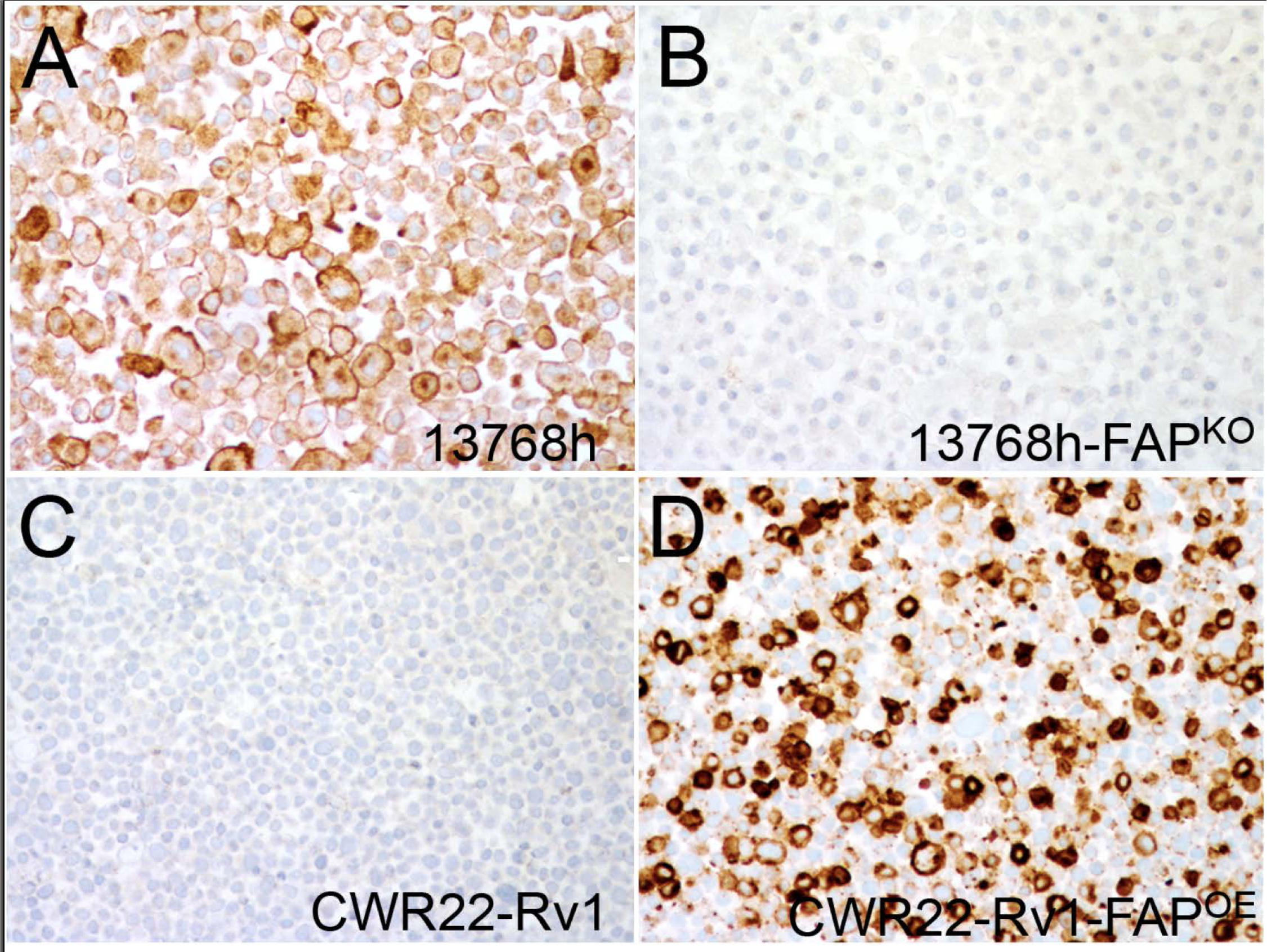
Analytic validation of FAP IHC assay. **(A)** hPrCSC-44c cells were immortalized from hPrCSC-44c cells that are known to be positive for FAP mRNA and protein and showed robust positive staining. (**B)** Targeted disruption of FAP using CRISPR/Cas9 resulted in an absence of IHC staining. **(C)** EnzaR-CWR22Rv1 prostate cancer cells are known to be negative for FAP and stained negatively prior to transfection, but were strongly positive following transfection with a FAP cDNA clone **(D)**.

To evaluate FAP expression in prostate tissues qualitatively, quantitatively and spatially, we developed a multiplex IHC assay using an iterative chromogenic approach (**Fig. 2**; **Supplemental Table 1**).^33,41^ An advantage of this iterative method is that each round of IHC is performed as a single-plex stain which facilitates quality control assessments for each individual marker before the next round of staining (**Fig. 2**). Each whole slide image is then subjected to color deconvolution and the pseudo-colored whole slide images from each staining round are fused to generate a multiplex pseudo-multicolored image^33^ (**Fig. 2**). To evaluate FAP expression by tissue compartment (*e.g.*, stroma versus epithelial), we used CK8, which stains all prostatic epithelial cells (benign and cancerous).^33^ To aid in the determination of benign versus cancerous regions, as well as intraductal carcinoma, the panel also includes the basal cell marker, p63, which is expressed in basal cells outlining all benign glands (including PIA) as well as those containing intraductal carcinoma and prostatic intraepithelial neoplasia (PIN).^33^ We included CD163 for M2 macrophages^42^ and CD45 for total immune cells.

**Figure 2.**
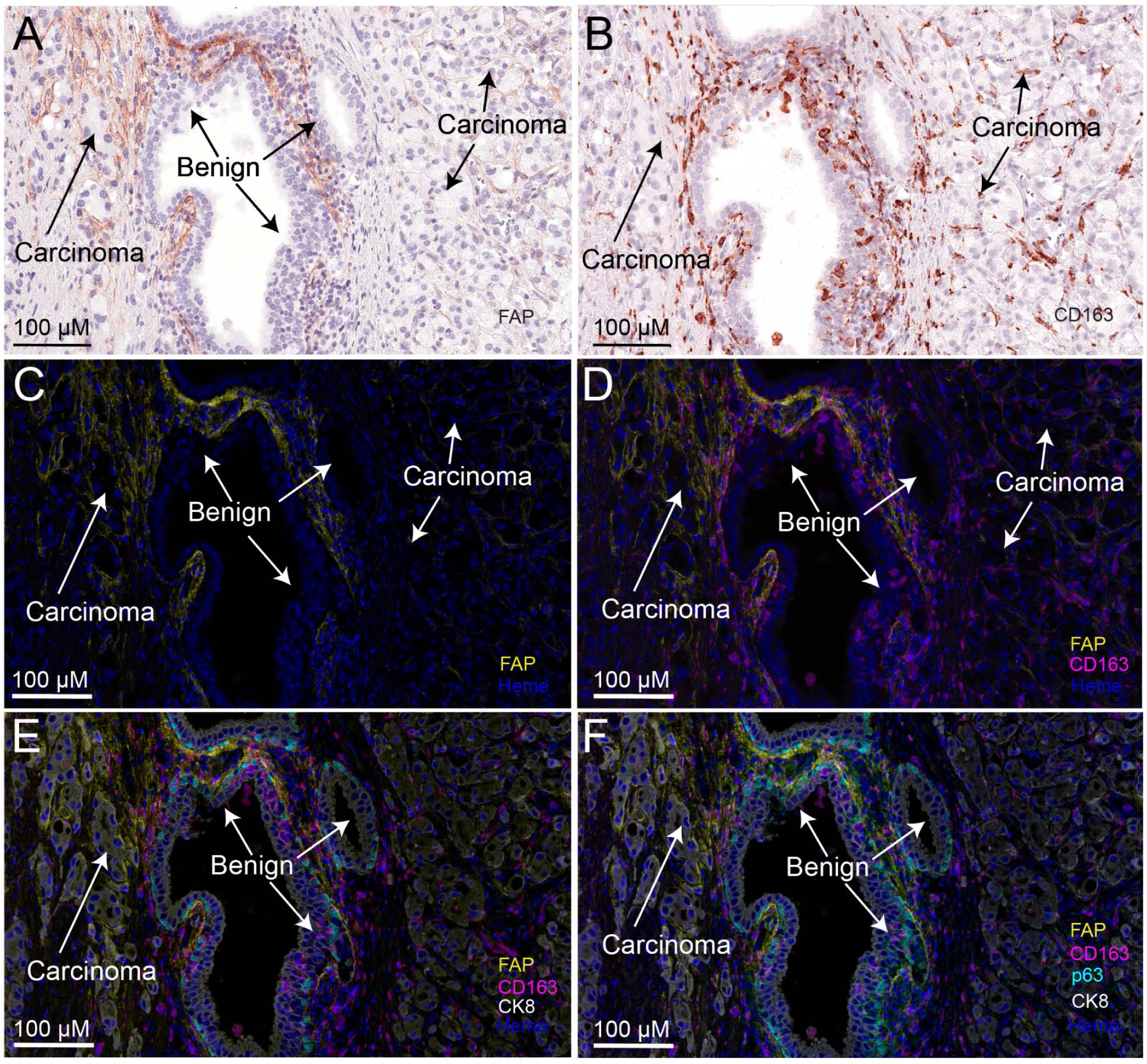
Multiplex Chromogenic IHC assay showing FAP expression in tumor region. FAP expression is restricted to the tumor stromal compartment. **A)** View of chromogenic stain of FAP in a region of a tumor in which malignant glands surround a benign acinus in the center. **B)** Iterative chromogenic stain on the same slide stained for CD163 IHC staining (M2 macrophages) in the same area described in **A. C)** Pseudocolored deconvoluted image of **A** showing FAP (yellow) and nuclei (blue). **D)** Same view as in **C** but with the addition of CD163 in fuchsia. **E)** Same view adding CK8 in white and p63 (basal cells) in cyan. **F)** Same view adding CD45 (pan immune cell marker) in green.

### FAP in Normal Appearing Regions and PIA

In regions of normal appearing prostatic tissue (without inflammation or atrophy) there was generally a complete absence of FAP staining (**Fig. 3A-B**). Regions of PIA, by contrast, often showed abundant, albeit heterogeneous, FAP staining in the stromal compartment (**Fig. 3D and F**). In some regions of PIA lesions, smooth muscle cells, which occupy the majority of the normal prostatic stroma area, were replaced by fibrosis (**Fig. 3E-F**), and these and surrounding areas generally showed high levels of FAP staining (**Fig. 3F**). PIA lesions often contain corpora amylacea, and often the most intense FAP staining was present in the stromal regions directly abutting these structures (**Fig. 3C-D and Fig. 5B**)

**Figure 3.**
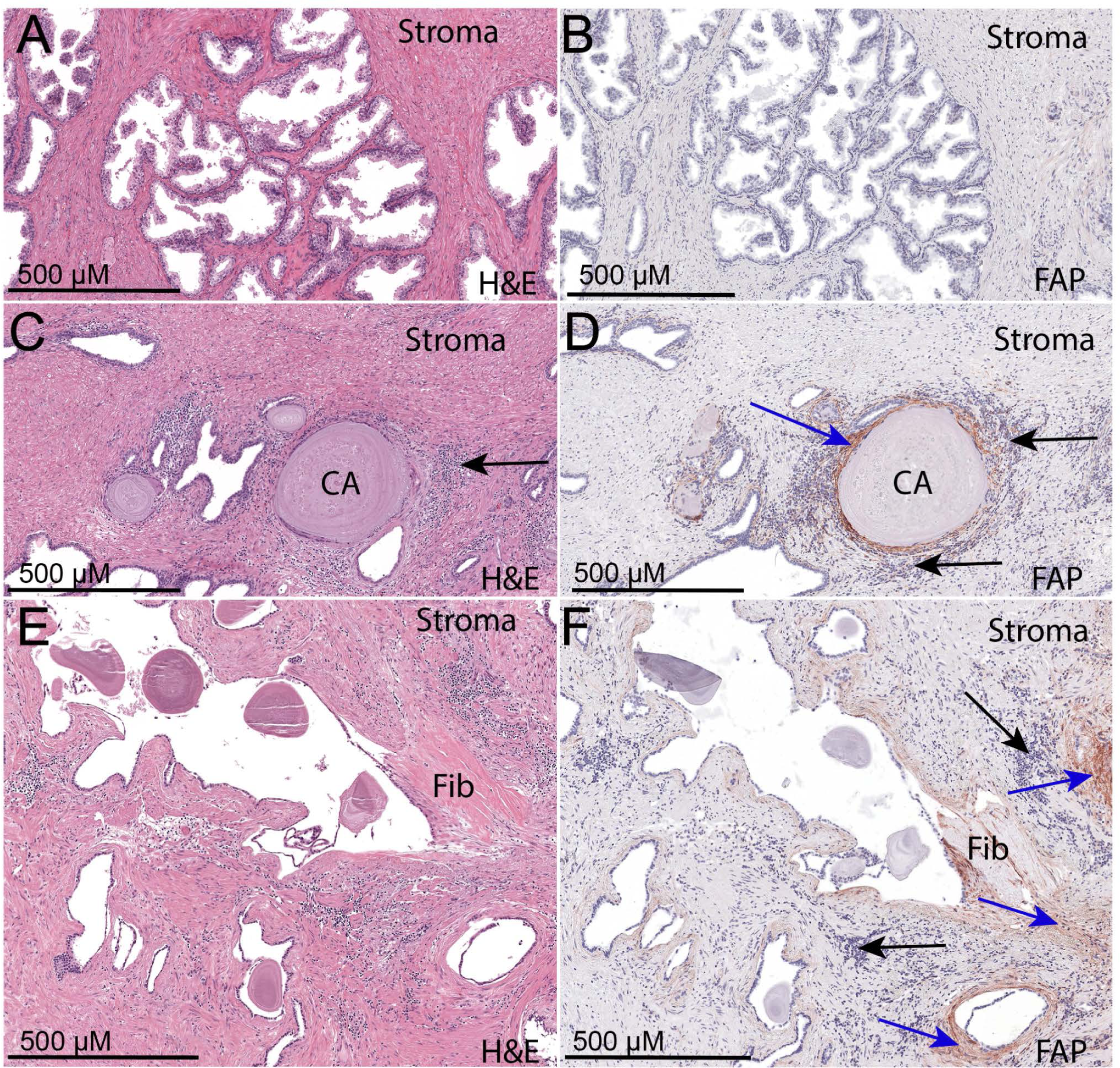
FAP IHC staining in benign prostate regions. **(A-B)** Normal prostatic epithelium and stroma show absence of staining for FAP protein. **(C-D)** Strong positive staining for FAP is present in the stromal compartment of a PIA lesion showing mild to moderate chronic inflammation and corpora amylacea (CA). Blue arrow indicates positive FAP staining. Black arrows indicate mononuclear inflammatory cells in the stroma. **(E-F)** H&E staining shows a region of fibrosis (Fib) surrounding an atrophic gland in another region of PIA. Blue arrows indicate positive areas of FAP staining in the stroma. Black arrows indicate mononuclear immune cell infiltrates.

### FAP in Prostatic Adenocarcinoma

In primary adenocarcinoma lesions, FAP staining was positive in all cases (N=56). Except for very focal staining in a small fraction of tumor cells in a single case, FAP was exclusively localized to the stromal compartment. Strikingly, in most cases, the extent and intensity of FAP staining was highly heterogeneous, often with some regions showing very high levels of staining and others containing much lower levels (**Fig. 4**).

**Figure 4.**
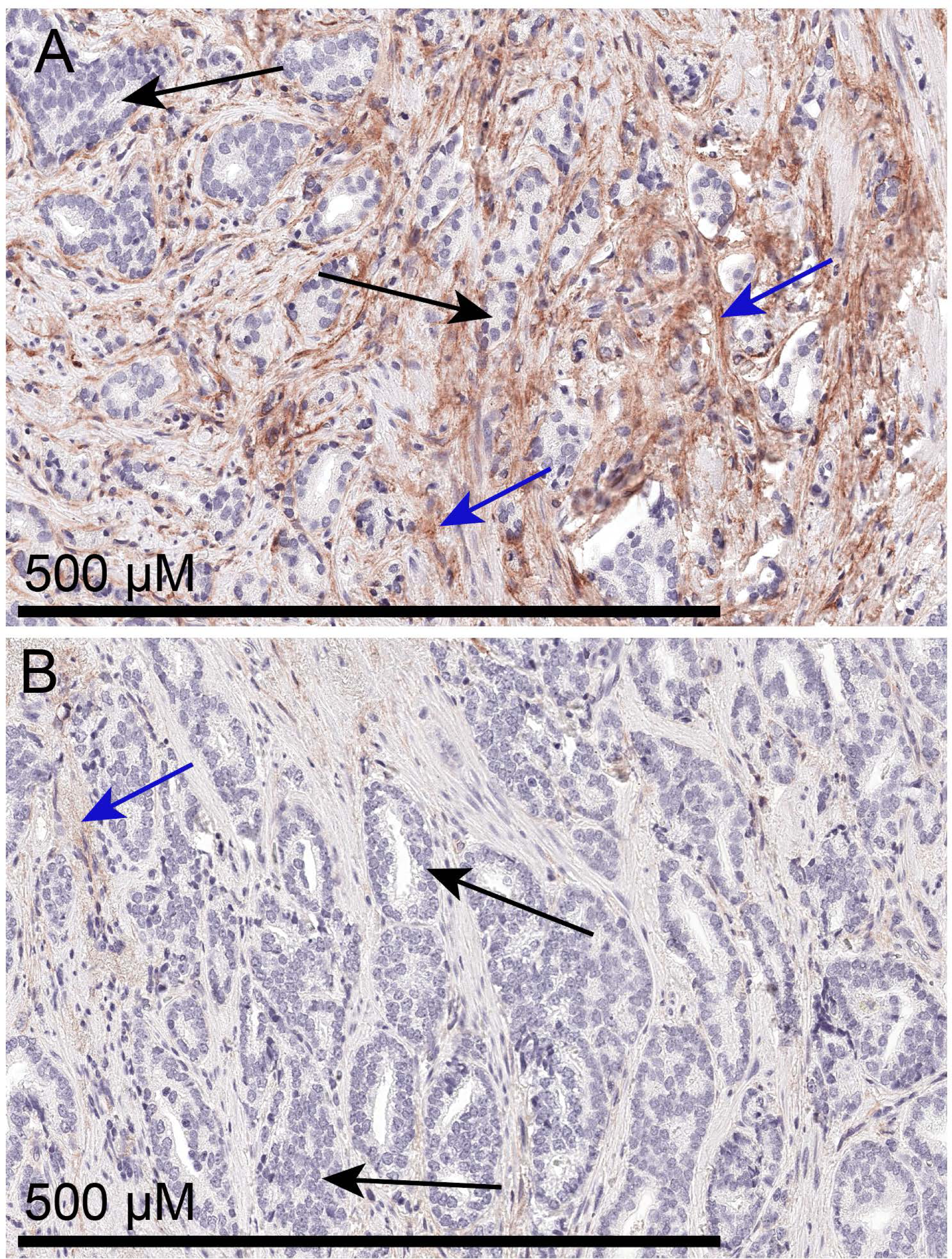
Marked heterogeneity of FAP in prostatic adenocarcinoma. **(A)** Region of invasive adenocarcinoma showing high levels of FAP staining in the tumor stromal compartment. **(B)** Another region from the same tumor has much less FAP staining. Black arrows indicate epithelial tumor cells and blue arrows indicate stromal regions staining positively for FAP.

**Figure 5.**
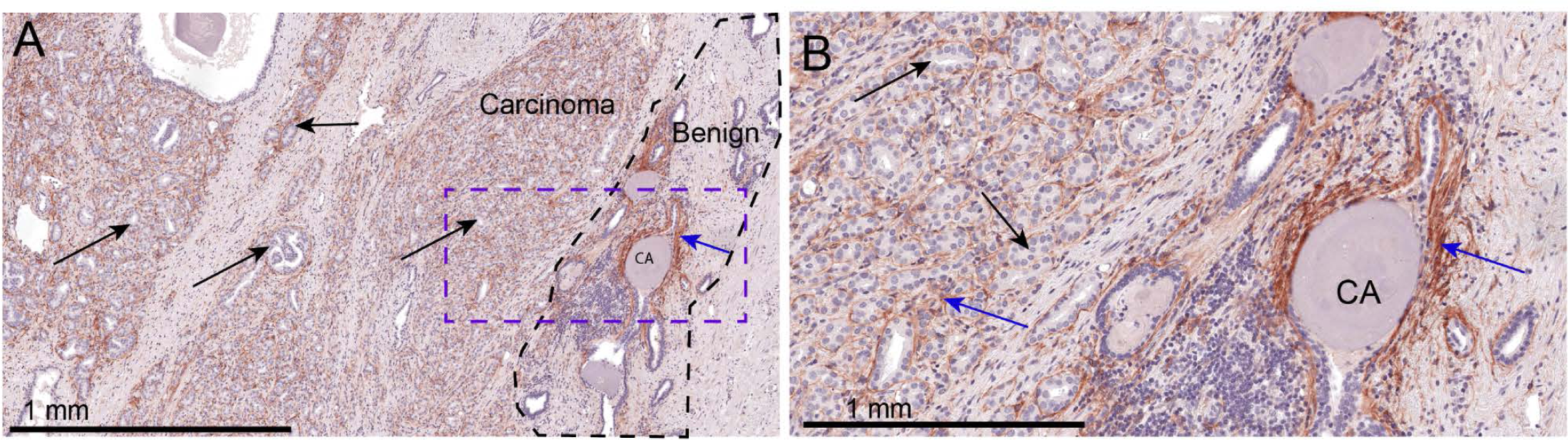
Strongest FAP staining occurs near benign glands even more so than nearby positively staining tumor. **(A)** Low power view showing region of invasive adenocarcinoma (left) directly adjacent to benign PIA glands and stroma (region within dashed black line). Strongest staining for FAP (blue arrow) is around benign gland that contains corpora amylacea (CA). Black arrows indicate tumor epithelial cells. Blue arrows indicate positive FAP staining in stroma. **(B)** Higher power view of region in **A** outlined with the blue dashed line.

Primary prostatic adenocarcinomas often contain malignant glands infiltrating between and around nearby benign glands. In tumors characterized by this growth pattern, the highest levels of FAP staining generally occurred directly surrounding benign atrophic glands of PIA (**Fig. 5).**

In carcinoma lesions with cribriform or intraductal architecture, FAP staining was not present within the tumor cell epithelial nodules, but was commonly present, often at high levels, in the stroma directly abutting the perimeter of these structures (**Fig. 6),** which is similar to that seen recently for FAP mRNA localization.^15^

**Figure 6.**
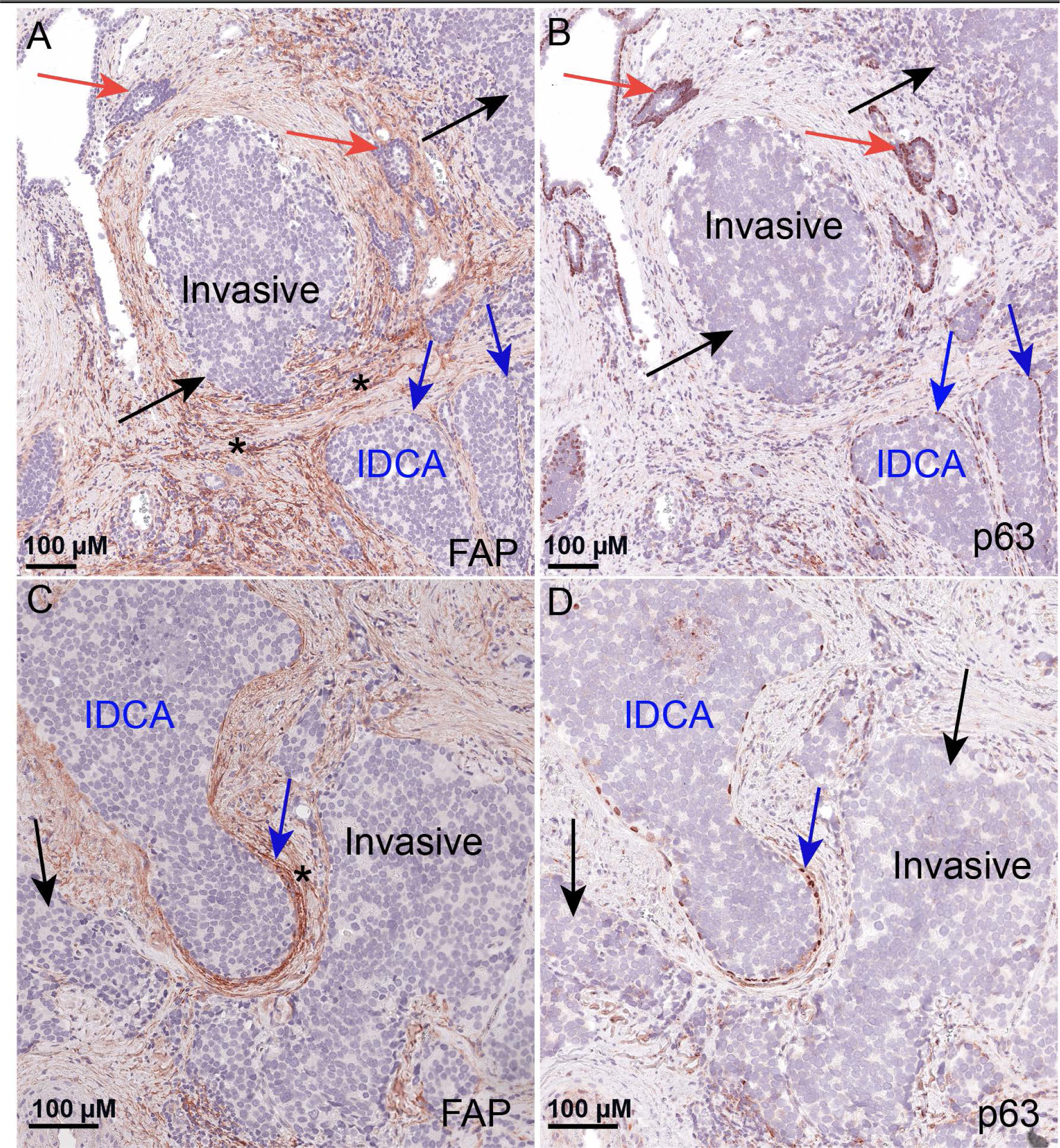
High levels of FAP occur around cribriform and intraductal lesions. Images show medium power view of chromogenic FAP staining (**A and C**) and p63 (**B and D**, highlighting nuclei of basal cells) of a high grade lesion with invasive adenocarcinoma (black arrows - invasive) and intraductal carcinoma (blue arrows - IDCA).Red arrows shows benign glands. Note strong FAP staining (*) in the stroma that is localized adjacent to epithelial tumor cell clusters.

### Quantitative Image Analysis Comparing Normal PIA and Carcinoma Regions

We next quantified the area of FAP staining in the different tissue types encompassing normal appearing regions, PIA, and invasive adenocarcinomas. For each of these tissue types, we determined the percentage of area occupied by FAP staining in relation to total tissue area as well as to stromal area. To quantify the epithelial and stromal tissue areas separately (as well as the total tissue area), we implemented a random forest classifier using keratin 8 IHC staining as a machine learning training guide^33^ (**Fig. 7**). We calculated the percentage area of FAP staining (FAP area fraction) in randomly selected areas from each tissue type where each tissue type was present on the same slides (N=5 matched patient pairs). **Fig. 8 and Table 2** show that the FAP area fraction was extremely low or absent in normal regions, but was increased in regions of PIA (**Fig. 8** shows one spot on each graph per patient after taking the median values and **Supplemental Fig. 1** shows all data points for all individual regions examined). In carcinoma, the FAP area fraction was also increased, compared with normal appearing regions, albeit it was not as high as in PIA. Exploring mRNA expression data from a recent study (The JHU LCM Prostatectomy Dataset) from our group using laser capture microdissection,^43^ FAP was overexpressed in tumor regions compared with matched normal regions (**Supplemental Fig. 2**). This provides additional evidence for FAP overexpression in primary prostatic adenocarcinoma.

**Figure 7.**
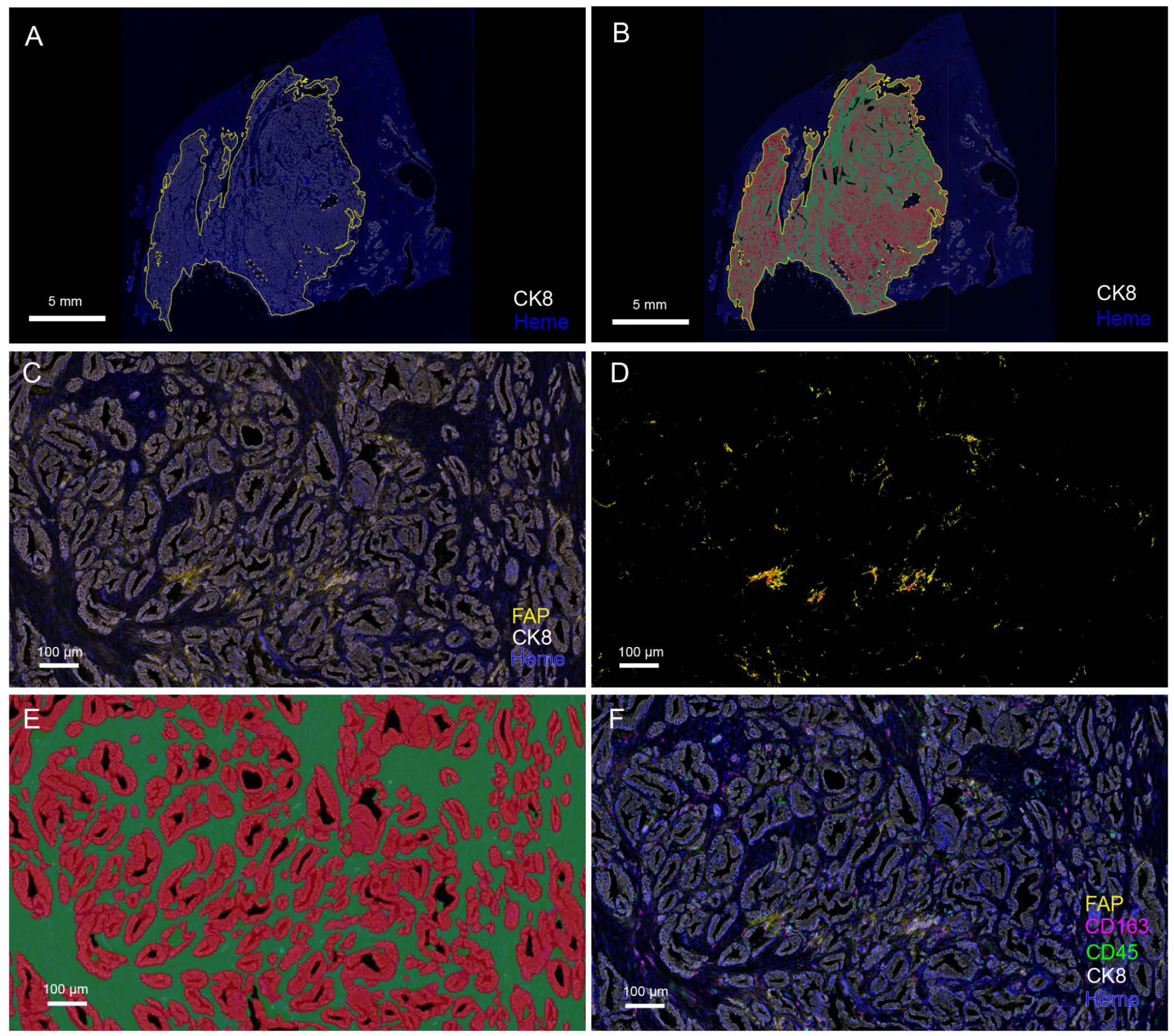
Multiplex chromogenic IHC assay and image analysis pipeline showing FAP area quantification. FAP expression is restricted to the tumor stromal compartment. **A)** low power view of whole slide with area of tumor demarcated in yellow. **B**) Use of Random Forest Classifier to detect tumor epithelium (red), stroma (green), and gland lumens (black). **C**) higher power view of tumor with FAP (yellow), epithelium (white), and nuclei (blue). **D**) Automated image analysis detection of FAP for area quantification (red is strong intensity, orange is medium and yellow is low intensity). E) Same region as C and D showing Random Forest Classifier as in **B. F**) Markers used for panel 1 (CD45 (green-all immune cells), CD163 in fuchsia (M2 macrophages), FAP (yellow) and keratin 8 (white).

**Figure 8.**
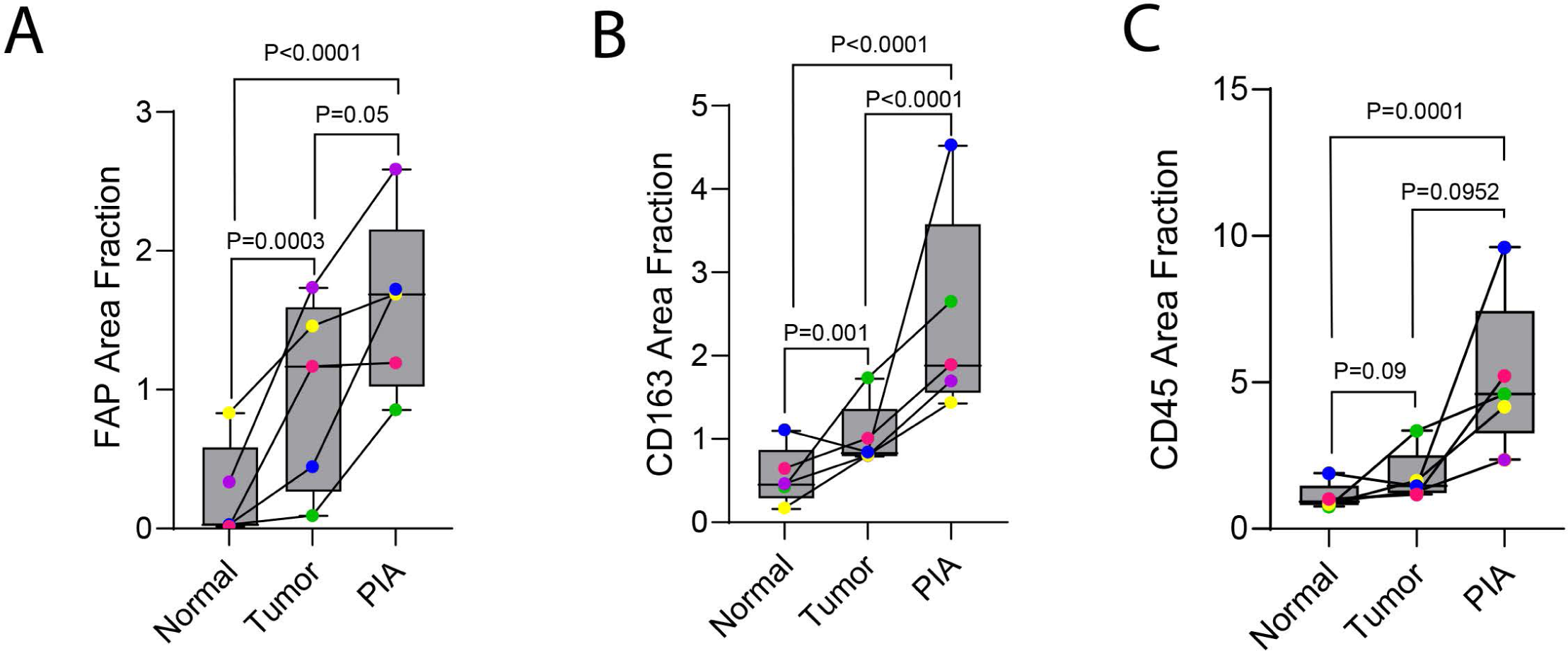
FAP area fraction and paired analyses from different regions (normal, PIA and tumor) from matched patients. **A-C)**. Quantification using the median value. Each point on the graph represents the median value from multiple regions from a given patient. Each color represents matched regions from the same patient.

**Table 2.** FAP area fraction (high and low thresholds) in random selected areas of normal appearing epithelium, PIA and Tumor.

|  |  | Total FAP |  |  | High Threshold FAP |  |  |
| --- | --- | --- | --- | --- | --- | --- | --- |
| Case |  | Normal | Carcinoma | PIA | Normal | Carcinoma | PIA |
| 1 | median | 0.02853 | 0.093663 | 0.856993 | 0.005911 | 0.014668 | 0.23805 |
|  | mean | 0.032737 | 0.1202954 | 1.491368 | 0.0058593 | 0.014329 | 0.423866 |
|  | SD | 0.13157 | 0.668347 | 1.519038 | 0.005543 | 0.0085454 | 0.4955947 |
| 2 | median | 0.337393 | 1.735095 | 2.589456 | 0.032995 | 0.215894 | 0.504221 |
|  | mean | 0.2862057 | 1.488986 | 3.330993 | 0.031131 | 0.2026046 | 0.9165356 |
|  | SD | 0.1046561 | 1.078719 | 2.67103 | 0.0072939 | 0.1717262 | 0.8783959 |
| 3 | median | 0.833499 | 1.458605 | 1.68821 | 0.086693 | 0.136588 | 0.264828 |
|  | mean | 0.8581937 | 1.356314 | 1.755053 | 0.935353 | 0.1619312 | 0.2577532 |
|  | SD | 0.0473302 | 0.682906 | 0.7428295 | 0.0184263 | 0.1202342 | 0.134802 |
| 4 | median | 0.29669 | 0.445324 | 1.722643 | 0.002331 | 0.062162 | 0.338115 |
|  | mean | 0.891427 | 1.285409 | 3.34904 | 0.0082647 | 0.2323456 | 1.460107 |
|  | SD | 0.1094155 | 1.65396 | 4.041396 | 0.0108505 | 0.3471015 | 2.328966 |
| 5 | median | 0.01702 | 1.168963 | 1.193922 | 0.002163 | 0.18999 | 0.161467 |
|  | mean | 0.17037 | 2.186287 | 1.396483 | 0.0025597 | 1.036728 | 0.4118354 |
|  | SD | 0.113835 | 3.183496 | 1.657962 | 0.0021773 | 1.328368 | 0.6160603 |

Next, we quantified the area fraction of CD163 positive macrophages and total CD45 positive total immune cells in these same regions. As expected, the area of M2 macrophages and overall immune cells was elevated in PIA as compared with normal appearing regions (**Fig. 8; Supplemental Fig. 1**). Also, both CD163 positive macrophages and total CD45 positive immune cells were elevated in the regions of adenocarcinoma – as compared with normal regions – and were lower than in PIA (**Fig. 8; Supplemental Fig. 1**).

### Quantitative Analysis of FAP in Carcinoma in Whole Tumor Regions

We next assessed the FAP area fraction in overall tumor regions for all 56 invasive carcinomas. For this, we annotated and quantified FAP staining within the entire tumor region present on standard whole slides for each case (**Fig. 7** shows an example). We examined FAP levels in relation to Gleason grade groups, pathological stage, and other morphological features (Figure 9, **Tables 3a, 3b and 3c**).

**Figure 9.**
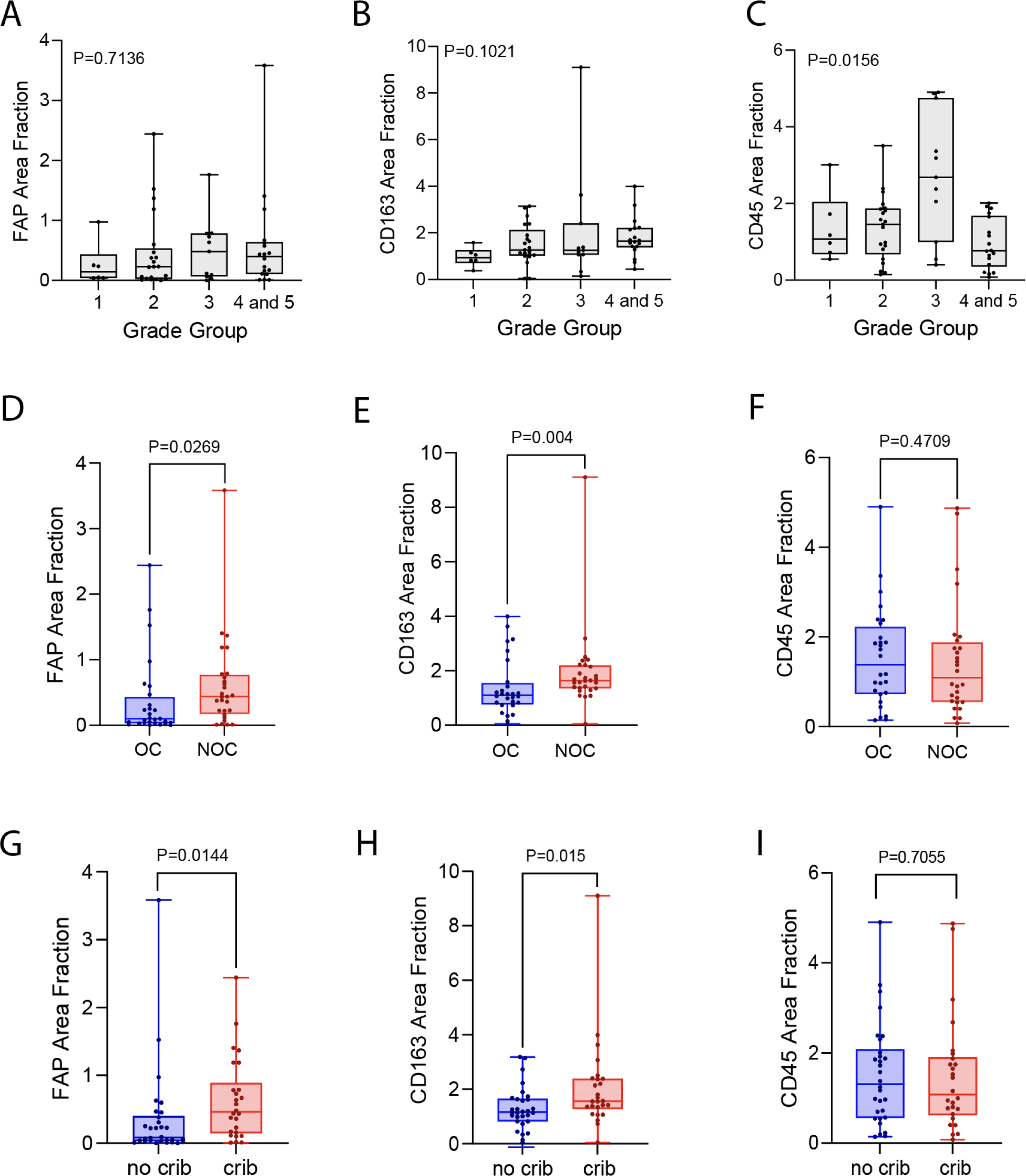
Quantitative analysis of FAP, CD163 and CD45 in carcinoma by grade, stage and the presence of cribriform glands. **A-C)** Quantification by grade group. **D-F)** Quantification by pathological stage at prostatectomy (organ confined versus non organ confined status). **G-I)**, Quantification by the presence of cribriform glands in carcinoma.

FAP expression in Grade Group 1 tumors (Gleason score 3+3=6) was generally quite low with one exception (**Table 3a**, **Fig. 9A**). There was a trend of increasing FAP protein levels with increasing grade groups, but this trend was relatively weak and not statistically significant (**Table 3a**, **Fig. 9A**). A similar trend was seen with CD163 positive macrophage area (**Table 3a**, **Fig. 9B**).

**Table 3a.** Associations between variables (P values shown).

| Variable |  | Grade |  |  | T2 vs > T2 |  |  | N1 vs No N1 |  |  | Tb3 vs <T3b |  |  |
| --- | --- | --- | --- | --- | --- | --- | --- | --- | --- | --- | --- | --- | --- |
|  |  | GG1 | all others | P | T2 | >T2 | P | no N1 | N1 | P | <T3b | T3b | P |
| Tumor FAP Total | Median | 1.21 | 1.71 | 0.44 | 2.36 | 3.39 | 0.09 | 1.69 | 4.65 | 0.05 | 1.73 | 3.72 | 0.13 |
|  | Range | 3.49 | 8.14 |  | 9.63 | 13.1 |  | 8.16 | 10.8 |  | 8.16 | 11.22 |  |
|  | SD | 1.36 | 2.32 |  | 2.99 | 3.01 |  | 1.89 | 5.01 |  | 1.91 | 4.75 |  |
| FAP area fraction | Median | 0.26 | 0.51 | 0.52 | 0.37 | 0.6 | 0.03 | 0.42 | 1.29 | 0.07 | 0.42 | 1.17 | 0.04 |
|  | Range | 0.94 | 3.58 |  | 2.44 | 3.57 |  | 2.44 | 3.22 |  | 2.44 | 3.42 |  |
|  | SD | 0.36 | 0.69 |  | 0.6 | 0.71 |  | 0.53 | 1.54 |  | 0.53 | 1.4 |  |
| FAP stroma total | Median | 0.38 | 0.77 | 0.33 | 2.36 | 3.39 | 0.09 | 2.59 | 6.59 | 0.04 | 2.63 | 5.4 | 0.07 |
|  | Range | 1.38 | 3.96 |  | 9.63 | 13.1 |  | 9.63 | 10.78 |  | 9.63 | 11.9 |  |
|  | SD | 0.53 | 0.89 |  | 2.99 | 3.01 |  | 2.71 | 4.67 |  | 2.72 | 4.88 |  |
| FAP stroma fraction | Median | 1.73 | 3.02 | 0.37 | 0.54 | 0.92 | 0.02 | 0.65 | 1.83 | 0.04 | 0.64 | 1.65 | 0.02 |
|  | Range | 1.38 | 3.96 |  | 3.18 | 3.95 |  | 3.18 | 3.14 |  | 3.18 | 3.73 |  |
|  | SD | 5.09 | 13.16 |  | 0.84 | 0.86 |  | 0.76 | 1.43 |  | 0.76 | 1.38 |  |
| CD163 area fraction | Median | 0.97 | 1.78 | 0.04 | 1.37 | 1.97 | 0.004 | 1.61 | 2.29 | 0.06 | 1.62 | 2.01 | 0.06 |
|  | Range | 1.2 | 9.06 |  | 3.95 | 9.06 |  | 9.06 | 1.77 |  | 9.06 | 1.84 |  |
|  | SD | 0.4 | 1.37 |  | 1.05 | 1.52 |  | 1.35 | 0.73 |  | 1.37 | 0.72 |  |
| CD45 area fraction | Median | 1.36 | 1.51 | 0.89 | 1.54 | 1.46 | 0.47 | 1.54 | 0.89 | 0.27 | 1.56 | 0.84 | 0.13 |
|  | Range | 2.46 | 4.82 |  | 4.76 | 4.8 |  | 4.76 | 1.97 |  | 4.82 | 1.56 |  |
|  | SD | 0.9 | 1.21 |  | 1.11 | 1.27 |  | 1.19 | 0.94 |  | 1.21 | 0.64 |  |

**Table 3b.**
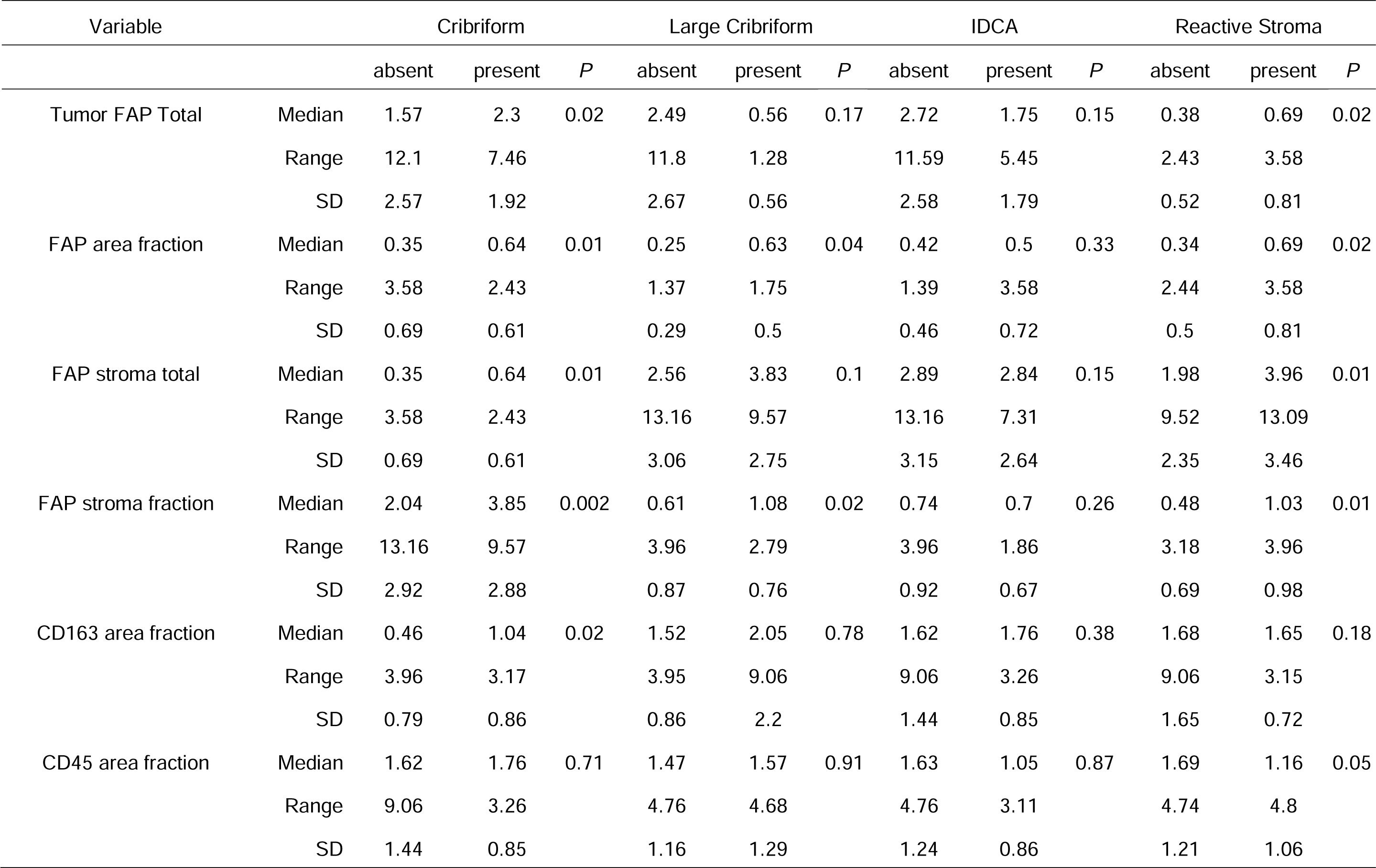
Associations between variables (P values shown).

| Variable |  | Cribriform |  |  | Large Cribriform |  |  | IDCA |  |  | Reactive Stroma |  |  |
| --- | --- | --- | --- | --- | --- | --- | --- | --- | --- | --- | --- | --- | --- |
|  |  | absent | present | *P* | absent | present | *P* | absent | present | *P* | absent | present | *P* |
| Tumor FAP Total | Median | 1.57 | 2.3 | 0.02 | 2.49 | 0.56 | 0.17 | 2.72 | 1.75 | 0.15 | 0.38 | 0.69 | 0.02 |
|  | Range | 12.1 | 7.46 |  | 11.8 | 1.28 |  | 11.59 | 5.45 |  | 2.43 | 3.58 |  |
|  | SD | 2.57 | 1.92 |  | 2.67 | 0.56 |  | 2.58 | 1.79 |  | 0.52 | 0.81 |  |
| FAP area fraction | Median | 0.35 | 0.64 | 0.01 | 0.25 | 0.63 | 0.04 | 0.42 | 0.5 | 0.33 | 0.34 | 0.69 | 0.02 |
|  | Range | 3.58 | 2.43 |  | 1.37 | 1.75 |  | 1.39 | 3.58 |  | 2.44 | 3.58 |  |
|  | SD | 0.69 | 0.61 |  | 0.29 | 0.5 |  | 0.46 | 0.72 |  | 0.5 | 0.81 |  |
| FAP stroma total | Median | 0.35 | 0.64 | 0.01 | 2.56 | 3.83 | 0.1 | 2.89 | 2.84 | 0.15 | 1.98 | 3.96 | 0.01 |
|  | Range | 3.58 | 2.43 |  | 13.16 | 9.57 |  | 13.16 | 7.31 |  | 9.52 | 13.09 |  |
|  | SD | 0.69 | 0.61 |  | 3.06 | 2.75 |  | 3.15 | 2.64 |  | 2.35 | 3.46 |  |
| FAP stroma fraction | Median | 2.04 | 3.85 | 0.002 | 0.61 | 1.08 | 0.02 | 0.74 | 0.7 | 0.26 | 0.48 | 1.03 | 0.01 |
|  | Range | 13.16 | 9.57 |  | 3.96 | 2.79 |  | 3.96 | 1.86 |  | 3.18 | 3.96 |  |
|  | SD | 2.92 | 2.88 |  | 0.87 | 0.76 |  | 0.92 | 0.67 |  | 0.69 | 0.98 |  |
| CD163 area fraction | Median | 0.46 | 1.04 | 0.02 | 1.52 | 2.05 | 0.78 | 1.62 | 1.76 | 0.38 | 1.68 | 1.65 | 0.18 |
|  | Range | 3.96 | 3.17 |  | 3.95 | 9.06 |  | 9.06 | 3.26 |  | 9.06 | 3.15 |  |
|  | SD | 0.79 | 0.86 |  | 0.86 | 2.2 |  | 1.44 | 0.85 |  | 1.65 | 0.72 |  |
| CD45 area fraction | Median | 1.62 | 1.76 | 0.71 | 1.47 | 1.57 | 0.91 | 1.63 | 1.05 | 0.87 | 1.69 | 1.16 | 0.05 |
|  | Range | 9.06 | 3.26 |  | 4.76 | 4.68 |  | 4.76 | 3.11 |  | 4.74 | 4.8 |  |
|  | SD | 1.44 | 0.85 |  | 1.16 | 1.29 |  | 1.24 | 0.86 |  | 1.21 | 1.06 |  |

**Table 3c.** Associations between variables (P values shown).

| Variable |  | PTEN Any Loss |  |  | PTEN Homogenous Loss |  |  | ERG status |  |  |
| --- | --- | --- | --- | --- | --- | --- | --- | --- | --- | --- |
|  |  | loss | retained | *P* | loss | retained | *P* | negative | positive | *P* |
| Tumor FAP Total | Median | 2.45 | 1.71 | 0.15 | 2.7 | 1.77 | 0.29 | 1.85 | 2 | 0.81 |
|  | Range | 5.41 | 12.1 |  | 5.41 | 12.1 |  | 8.16 | 12.06 |  |
|  | SD | 1.99 | 2.37 |  | 2.24 | 2.32 |  | 2.05 | 2.72 |  |
| FAP area fraction | Median | 0.6 | 0.44 | 0.18 | 0.67 | 0.45 | 0.31 | 0.44 | 0.56 | 0.7 |
|  | Range | 1.39 | 3.58 |  | 1.39 | 3.58 |  | 2.44 | 3.58 |  |
|  | SD | 0.52 | 0.69 |  | 0.58 | 0.67 |  | 0.57 | 0.81 |  |
| FAP stroma total | Median | 3.55 | 2.6 | 0.21 | 4.08 | 1.77 | 0.35 | 2.9 | 2 | 0.95 |
|  | Range | 7.27 | 13.16 |  | 7.27 | 12.1 |  | 9.63 | 12.06 |  |
|  | SD | 2.69 | 3.01 |  | 3.03 | 2.32 |  | 2.91 | 2.72 |  |
| FAP stroma fraction | Median | 0.89 | 0.66 | 0.21 | 1.05 | 0.67 | 0.36 | 0.68 | 0.81 | 0.77 |
|  | Range | 1.86 | 3.96 |  | 1.86 | 3.96 |  | 3.18 | 3.96 |  |
|  | SD | 0.7 | 0.87 |  | 0.78 | 0.84 |  | 0.8 | 0.97 |  |
| CD163 area fraction | Median | 2.38 | 1.53 | 0.58 | 3.42 | 1.53 | 0.4 | 1.54 | 1.84 | 0.83 |
|  | Range | 8.09 | 3.95 |  | 8.06 | 3.95 |  | 3.94 | 9.06 |  |
|  | SD | 2.59 | 0.93 |  | 3.82 | 0.9 |  | 0.92 | 1.83 |  |
| CD45 area fraction | Median | 1.3 | 1.49 | 0.33 | 1.53 | 1.46 | 0.4 | 1.59 | 1.34 | 0.23 |
|  | Range | 4.56 | 4.82 |  | 4.56 | 4.82 |  | 4.74 | 4.68 |  |
|  | SD | 1.44 | 1.13 |  | 2.16 | 1.1 |  | 1.16 | 1.22 |  |

In terms of pathological stage at prostatectomy, the FAP area fraction was higher in cases with non-organ confined disease (>=pT3a) versus organ confined (pT2) (**Fig. 9**) and in cases considered high stage (pT3b showing seminal vesicle invasion or Any N1 showing lymph node metastasis) versus lower stages (<=pT3a) (Table 3). The CD163 positive macrophage area fraction, but not CD45 area, was also increased in higher stage cases (**Table 3a**; **Fig. 9E-F**).

In terms of morphological characteristics not encompassed by Gleason grade groups, we recorded whether carcinoma lesions harbored any reactive stroma in the nearby H&E stained sections and observed a correlation between FAP area fraction and the presence of reactive stroma (**Supplementary Figure 3**; **Table 3b**). Cribriform architecture is known to be associated with adverse outcomes, independent of Gleason scoring. Furthermore, a recent study found increased FAP mRNA in cribriform prostatic carcinoma.^17^ 46.4% of cases showed any cribriform glands; 25% had large cribriform glands. There was increased FAP in cases with any cribriform glands and with large cribriform glands (P = 0.04 rank-sum). FAP expression was not, however, associated with the presence of intraductal carcinoma (N = 13 cases with intraductal carcinoma, P = 0.972, rank-sum).

Prior studies found that FAP mRNA levels correlated with the extent of CD163 mRNA (used as surrogate for M2 macrophage infiltration) across multiple tumor types,^44^ and FAP protein and CD163 protein correlated in primary prostatic adenocarcinomas.^16^ In the present study, there was a positive correlation between the FAP area fraction and the CD163 area fraction (**Fig. 10A**). This is despite the fact that there was not an association between FAP area fraction and the extent of infiltration by total CD45 positive immune cells (**Fig. 10B**). There was a trend towards increased CD163 area fraction and Grade Groups, although this was not significant (**Fig. 9B**). In terms of CD45 area fraction, there was an apparent increase in Grade group 3 tumors, yet a somewhat lower level in combined high grade (Grade group 4 and 5) lesions (**Fig. 9C**). We did not observe appreciable colocalization of FAP and CD163, indicating that intratumoral prostatic macrophages generally do not express FAP. Exploring mRNA expression data from the The JHU LCM Prostatectomy Dataset, as well as from a prior microarray study in prostate prostatic adenocarcinomas^45^ and from the TCGA, FAP mRNA correlated with CD163 mRNA in both studies (**Supplemental Fig. 4**).

**Figure 10.**
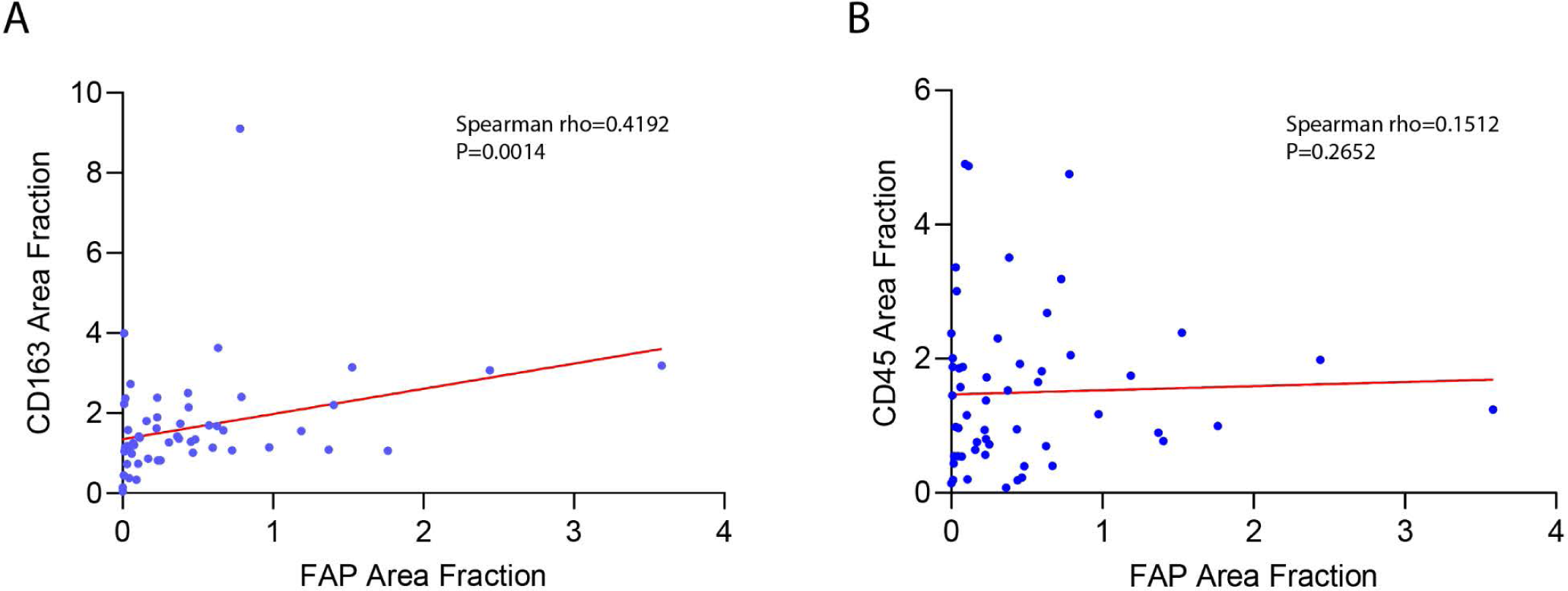
FAP area fraction in carcinoma correlates with CD163 area fraction, but not with CD45 area fraction.

### Relation of FAP Expression to Common Molecular Alterations

A prior study indicated that FAP expression was higher in prostate cancer cases with PTEN loss.^16^ Also, 25-50% of prostatic adenocarcinomas harbor ERG gene fusions, which denote a specific molecular subtype. We thus evaluated the level of FAP in relation to the status of PTEN and ERG as characterized by IHC.^33^ Although not statistically significant, there was a trend for increased area fraction of FAP staining in cases with PTEN loss as well as in cases that were ERG positive (**Supplemental Fig. 5**). The extent of CD163 positive area fraction was higher in cases with complete PTEN loss (**Supplemental Fig. 5**).

## Discussion

In this study, we report, for the first time, widespread and high levels of FAP expression within benign prostatic tissues in regions of PIA. Furthermore, our study is the first to use standard whole slides, which provide an advantage by markedly increasing the amount of tumor tissue examined for FAP expression, as compared with TMA spots. Using this approach, FAP protein was detected within the tumor microenvironment in all primary untreated prostatic adenocarcinomas examined. Using whole slides also revealed that FAP expression was generally highly heterogeneous within given tumors, which could in part explain prior discrepant results using TMAs. We validated our staining using genetic controls, which demonstrated very high specificity for FAP protein. Having null cell line controls allowed us to maximize our signal while keeping the noise extremely low, which provided high sensitivity for FAP protein detection. It is likely that this increased our ability to detect relatively low levels of FAP expression.

In addition to shedding light on the pathogenesis of prostate cancer development, the present results also have clinical implications. Molecular imaging and theranostic approaches targeting FAP using recently developed radiopharmaceuticals are proceeding rapidly and are considered highly promising as novel cancer imaging and therapeutic agents.^1,7–9^ Our findings suggest that future applications of molecular imaging targeting FAP, as well as FAP-targeted theranostics, should be considered worth pursuing in clinically localized prostate cancer; yet, at the same time, it is important to recognize that false positive imaging could occur if relatively large regions of FAP-positive PIA are present. Careful tissue mapping with registration between imaging results and histopathologic findings will be required to evaluate this accurately.

In non-neoplastic prostate tissues, we have proposed that in response to infectious agents (*e.g.*, bacteria, fungi, or viruses), inflammatory cell infiltrates, dietary toxicants, urine reflux, *etc.*, prostatic epithelial cells undergo cellular injury and cell death.^18–20^ Whether initiated by inflammation or otherwise, the cell injury/death would also be expected to further drive inflammation in a feed-forward loop.^19^ Further, to restore the barrier function, epithelial regeneration occurs via proliferation of intermediate epithelial cells, which are highly enriched in PIA lesions.^18–20,28^ Here, we provide the first evidence showing that regions of PIA often contain high levels of FAP protein in their stromal microenvironment. In fact, in many of the cases, the highest level of FAP expression occurred in PIA lesions. Along with morphological features showing overt fibrosis, these findings are consistent with the injury-regeneration model of PIA development and adds a new feature consisting of stromal injury. We postulate that this stromal injury occurs from damage to prostatic smooth muscle cells and fibroblasts, along with other elements such as small blood vessels and nerves, by the same mechanisms described above for epithelial cells.

These findings indicate that regions of PIA with increased FAP appear to represent a wound undergoing tissue remodeling, analogous to wounds of the skin and other body sites. The results also suggest a novel concept that those PIA lesions with increased FAP harbor a supportive microenvironment that increases their risk of progressing to PIN and/or adenocarcinoma. We hypothesize that, at times, PIN or early carcinoma lesions that develop in regions of high FAP may harbor a growth advantage due to the known function of FAP in promoting epithelial cell proliferation, new blood vessel formation, stromal invasion and immune suppression.^46,47^

In addition, the remodeling of the microenvironment and stroma around PIA lesions, which can be quite extensive throughout the peripheral zone and other regions of the prostate, could also be fertile ground for pre-existing infiltrating adenocarcinoma cells. For example, as they migrate into these regions, tumor cells may co-opt the pre-existing FAP for neoplastic progression. This suggests that some of what is considered “reactive stroma” in cancer, with cancer associated fibroblasts (CAFs), may actually represent pre-existing altered stroma occurring in the pre-cancer microenvironment that may be encountered by, and at times co-opted by, the tumor cells. However, in some lesions, prostatic carcinomas occupy relatively large areas with no intermingling of benign glands. In these lesions, it is likely that the extent of FAP present indicates new expression in reactive tumor stroma that was not necessarily preexisting. This could occur either through activation of tissue-resident fibroblasts leading to FAP upregulation, in part through TGF-β signaling ^12,48^ or through recruitment of mesenchymal precursors from systemic sources like the bone marrow.^35,49,50^

CAFs have been widely studied and are often considered part of the “reactive stroma,” which in prostate cancer has been associated with aggressive disease and poor outcomes.^51–54^ In the present study, we found that the extent of FAP expression in the tumor microenvironment correlates with the presence of reactive stroma, which has been previously reported in a small series of cases.^48^ This suggests that perhaps part of the association with reactive stroma with aggressive disease may relate to the presence of FAP overexpression. We also found that FAP levels were very low in grade group 1 tumors, but otherwise were not tightly associated with increasing grade group. However, FAP levels were associated with increased prostate cancer stage, which is an established marker of prostate cancer aggressiveness.

Prior studies have associated FAP with M2 macrophage infiltration.^16,44,55–57^ In the present study, the extent of FAP expression correlated with the extent of tumor infiltration by CD163 positive macrophages, which we verified were themselves higher in carcinoma than in normal appearing regions.^42^ Since these CD163-positive M2 macrophages are generally considered immunosuppressive, these findings are consistent with the hypothesis that FAP may contribute to immune suppression in the prostate tumor microenvironment.^12,35,58,59^ Interestingly, while FAP expression tended to occur in regions of inflammation, FAP expression did not correlate with overall CD45+ leukocyte density, suggesting some specificity of FAP and macrophage colocalization.

Our study has a number of limitations. First, we probed a relatively small number of cases and we do not have clinical follow-up on the cases studied. Also, we do not know the molecular mechanisms for FAP overexpression in human prostate stroma, benign or malignant. A number of cytokines and growth factors have been implicated in driving FAP expression. One prominent candidate for induction of FAP is TGF-β,^12,48,60,61^ although additional studies are required to determine if this mechanism is actively occurring in human prostate tissues.

Furthermore, while we have ruled out FAP expression by macrophages and CD45 positive immune cells in our study, we have not further characterized the presumptive fibroblasts expressing FAP. CAFs are phenotypically heterogeneous in terms of gene expression and function with recent studies also indicating a diversity of biological functions.^53,62,63^ Recent studies using single cell RNAseq have provided new information about heterogeneous subtypes of cell states of fibroblasts in the tumor microenvironment of the prostate.^17,32,64^ Given this, it will be important to better define their phenotypes and localization spatially and quantitatively in non-neoplastic and neoplastic prostatic lesions. Future studies including additional multiplex IHC panels and *in situ* transcriptomics can help provide clarity to these questions.

## Data Availability

All data produced in the present study are available upon reasonable request to the authors

## Acknowledgments

We thank Dr Charles J. Bieberich for careful reading of the manuscript and helpful insight. This work is supported by NIH/NCI U54 CA274370 (AMDM and SY) NIH/ National Cancer Institute (NCI) Specialized Programs of Research Excellence (SPORE) in Prostate Cancer grant P50CA58236 (AMDM), NIH/NCI grant U01 CA196390 (AMDM and SY), R01 CA255259-04(WNB), R01 CA279993-01A1 (WNB), U.S. Department of Defense Prostate Cancer Research Program (PCRP), W81XWH-18-2-0015 (AMDM), NIH/NIBIB P41 EB024495 (MP), The Johns Hopkins Sidney Kimmel Comprehensive Cancer Center Oncology Tissue Services Laboratory is supported by NIH/NCI grant P30 CA006973 and The Patrick C. Walsh Prostate Cancer Research Fund at Johns Hopkins (AMDM and SY).

---

**Supplemental Figure 1.**
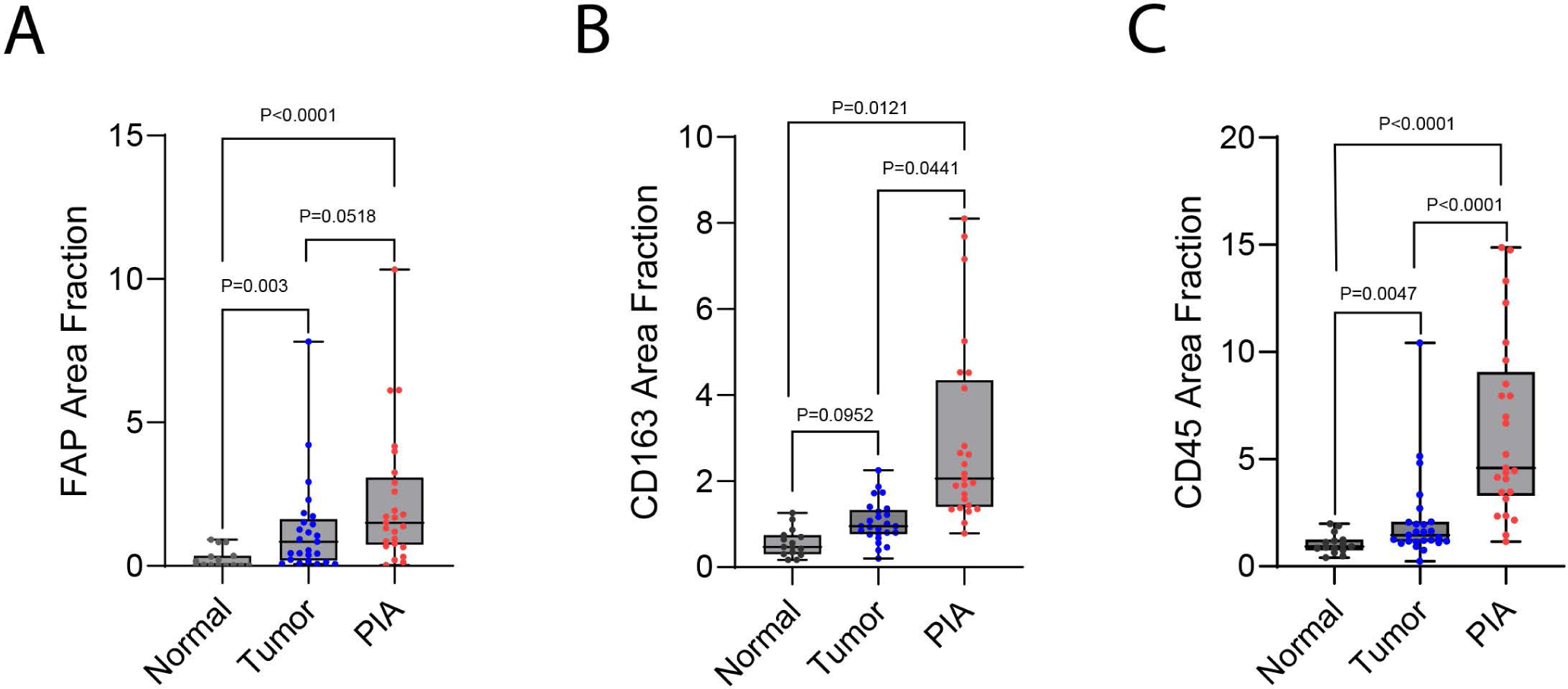
FAP area fraction from all data points used for data shown in Fig. 8. A-C) Each color dot represents the FAP area fraction (percentage of area occupied by FAP staining) in a given region of interest in that tissue type, including all areas (multiple areas per patient) from all 5 patients. There is a marked increase of FAP in tumor and PIA regions.

**Supplemental Figure 2.**
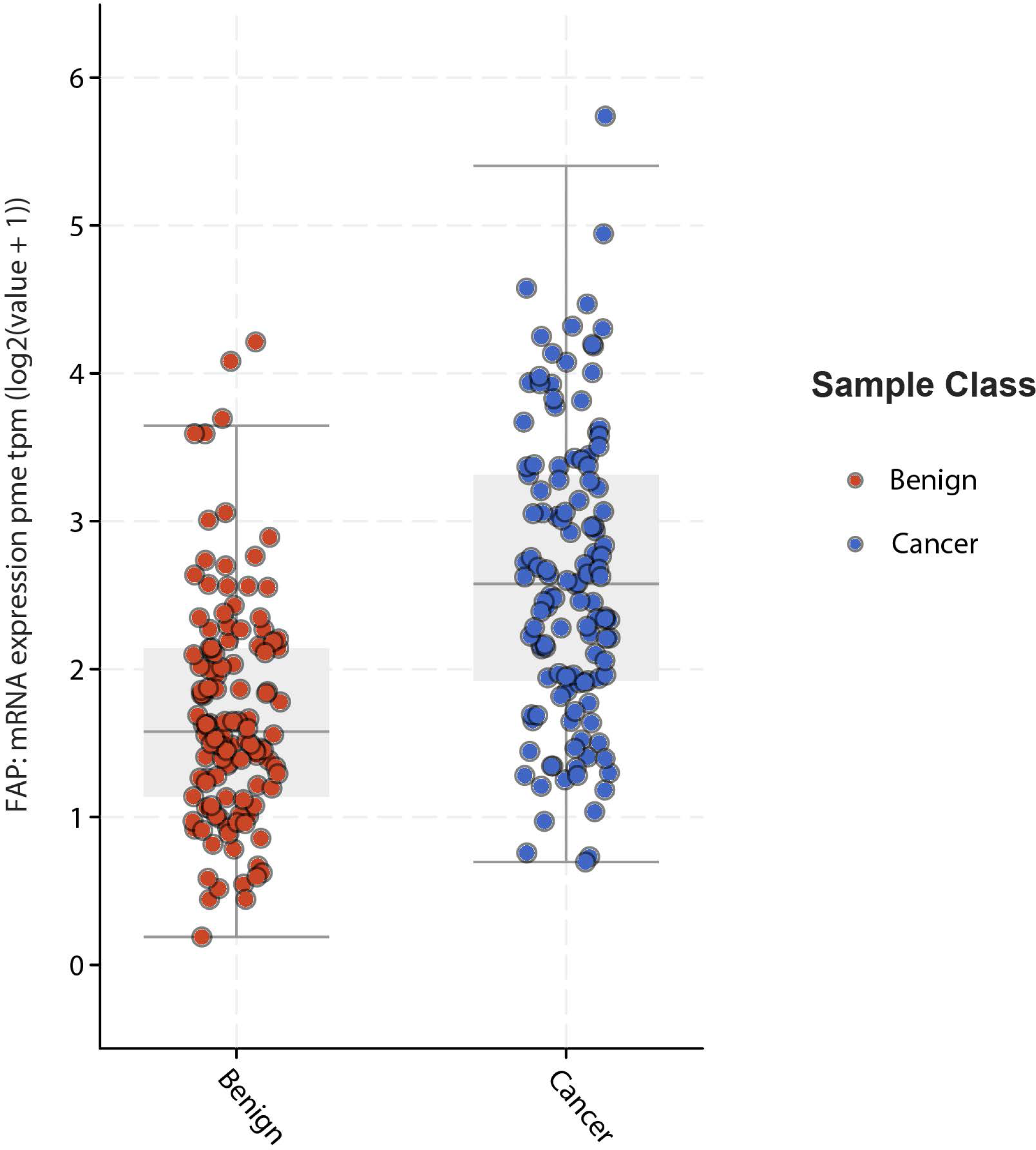
FAP mRNA expression data from The JHU LCM Prostatectomy Dataset^43^, showing its expression in benign and adenocarcinoma regions (N= 114 patients with cancer and 120 patients with benign). TPM represents transcripts per million data.

**Supplemental Figure 3.**
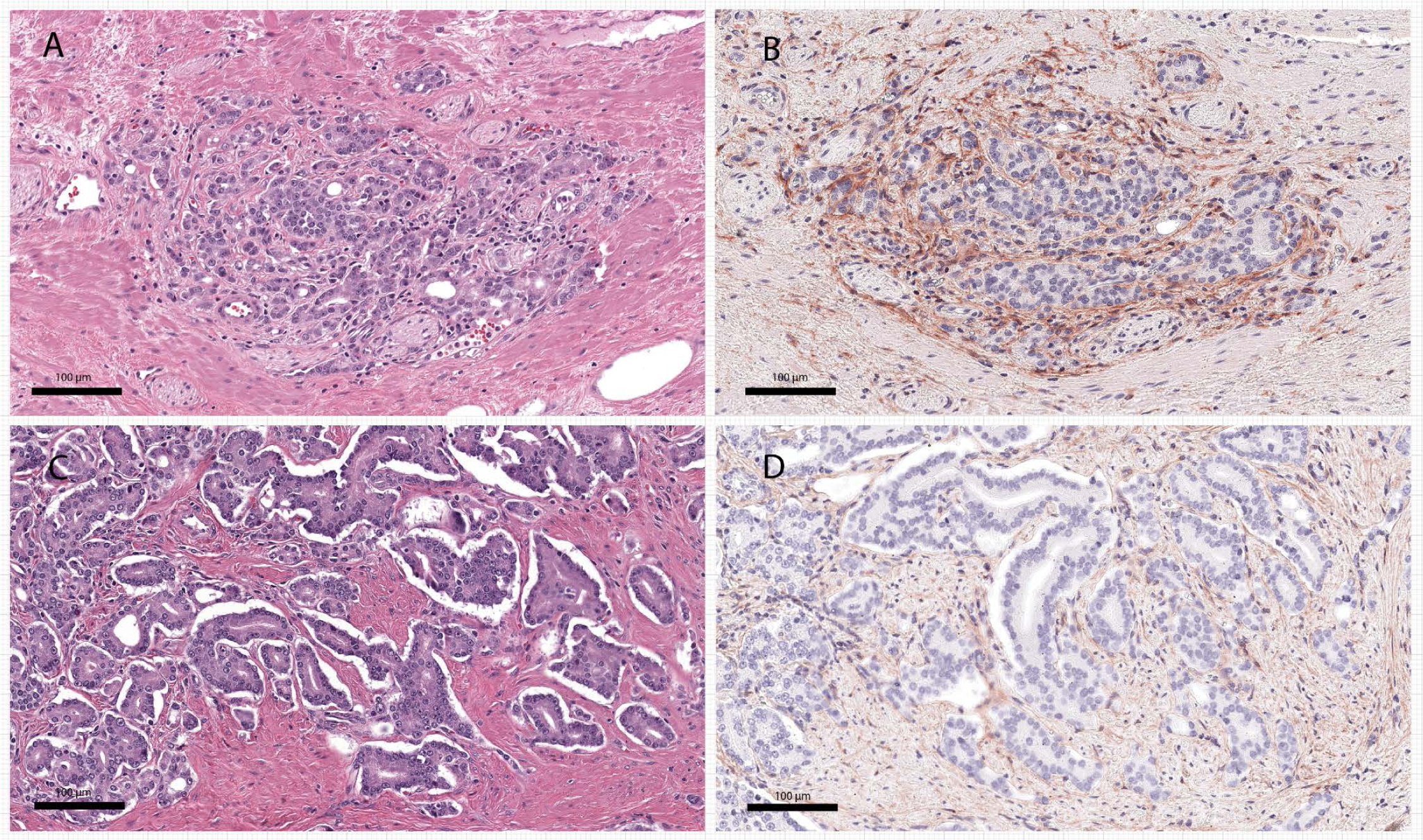
FAP expression in reactive stroma. **A)** H&E image of a region of prostatic adenocarcinoma showing reactive stroma between cancerous glands. **B)** FAP stained slide from an adjacent slide from the same region as in **A** showing intense staining in areas of reactive stroma. **C)** Same tumor in a different region, where there is no reactive stroma. **D)** FAP stained slide of the same view as **C**, showing low intense staining in areas lacking reactive stroma.

**Supplemental Figure 4.**
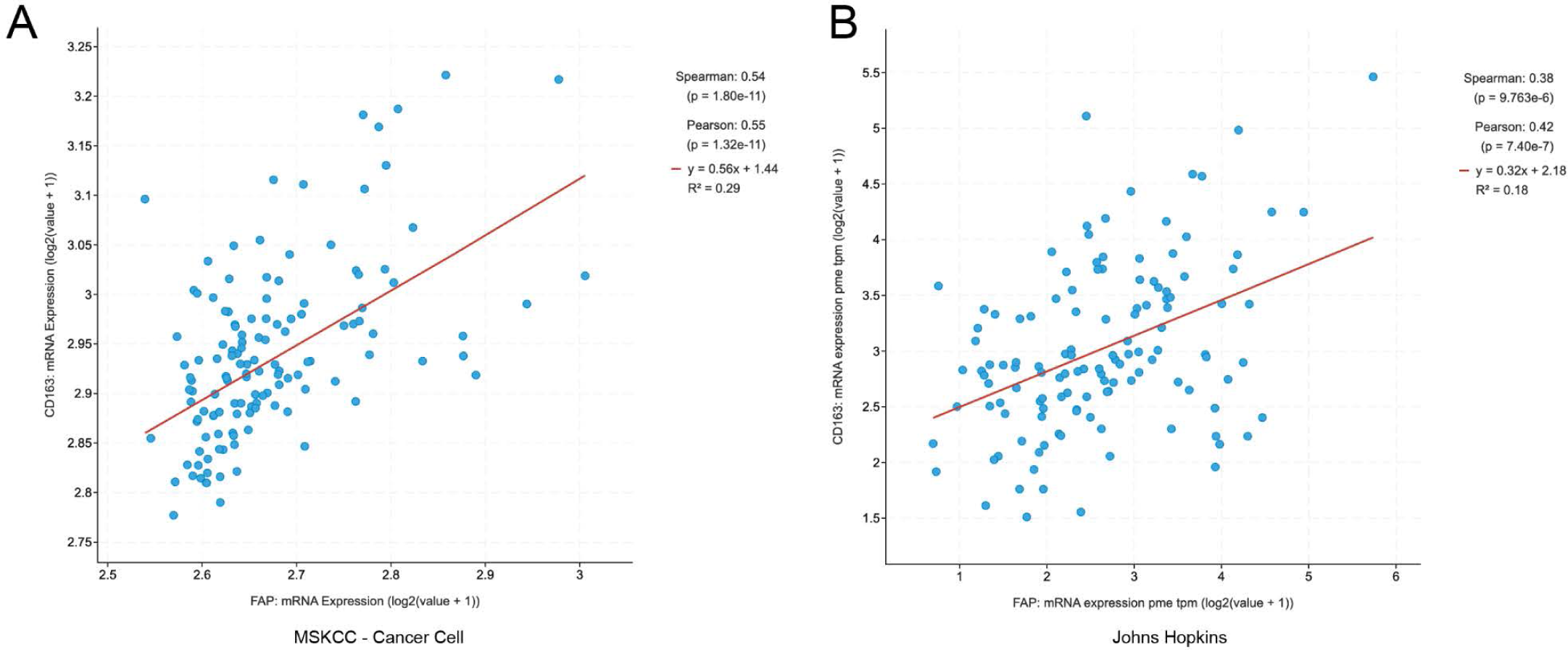
Correlation between FAP mRNA and CD163 mRNA expression from RNAseq datasets. **A**, Data from MSKCC *Cancer Cell*^45^ (N = 131 samples from primary tumors with mRNA). **B**, Data from Chen et al., Johns Hopkins, *JCI Insight*^43^ (N = 126 samples from 120 patients). **C**, Data from TCGA, Cell, 2015^65^ (N = 290 samples/patients).

**Supplemental Figure 5.**
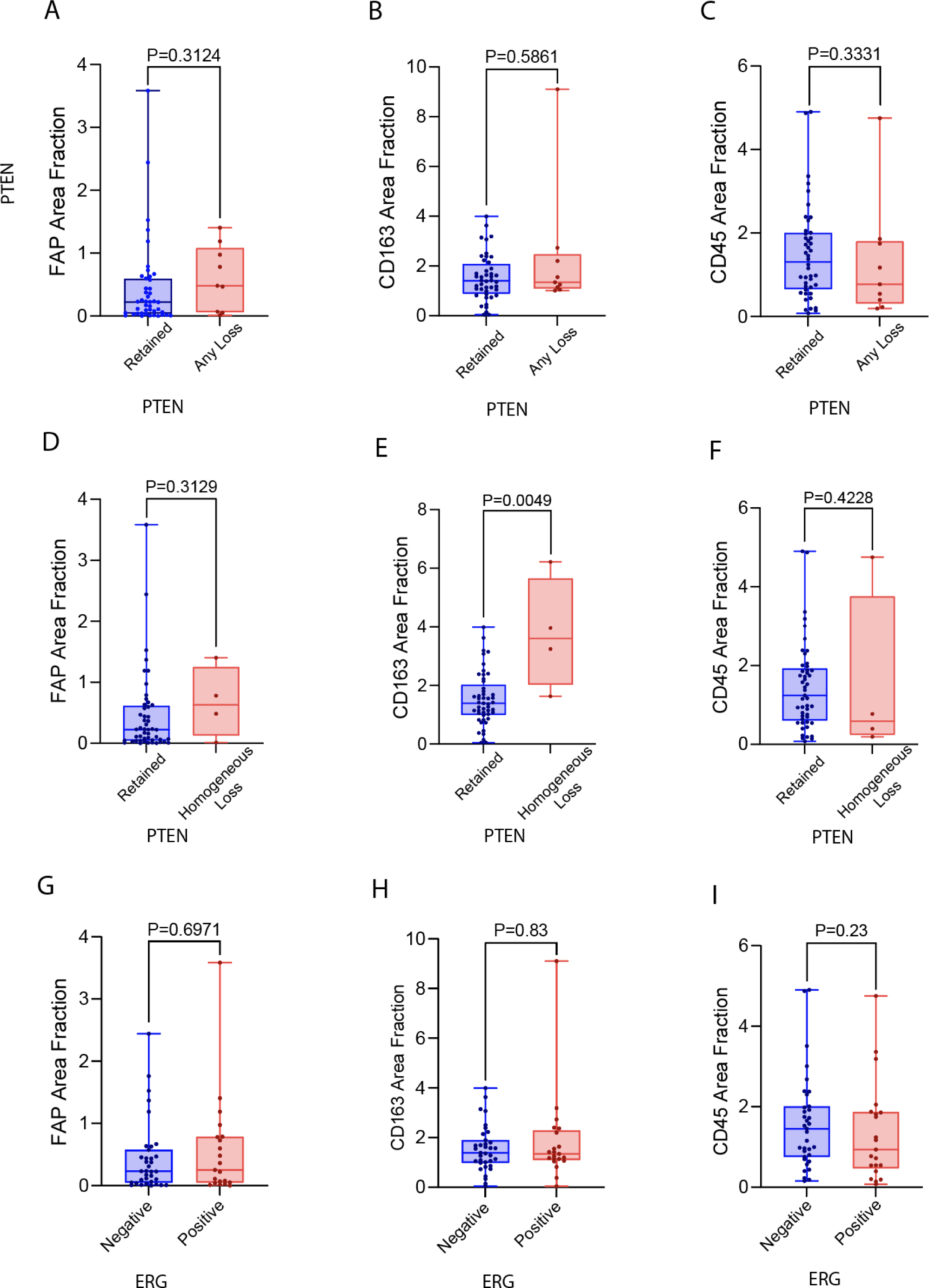
Quantitative analysis of FAP, CD163 and CD45 in relation to common molecular alterations in prostate cancer. **A-F)** Quantitative analysis of FAP, CD163, and CD45 area fraction in cases with any (heterogeneous) PTEN loss **(A-C)** or homogeneous PTEN loss **(D-F). G-I)** Quantitative analysis of FAP area fraction in cases in relation to ERG status.

**Supplemental Table 1.**
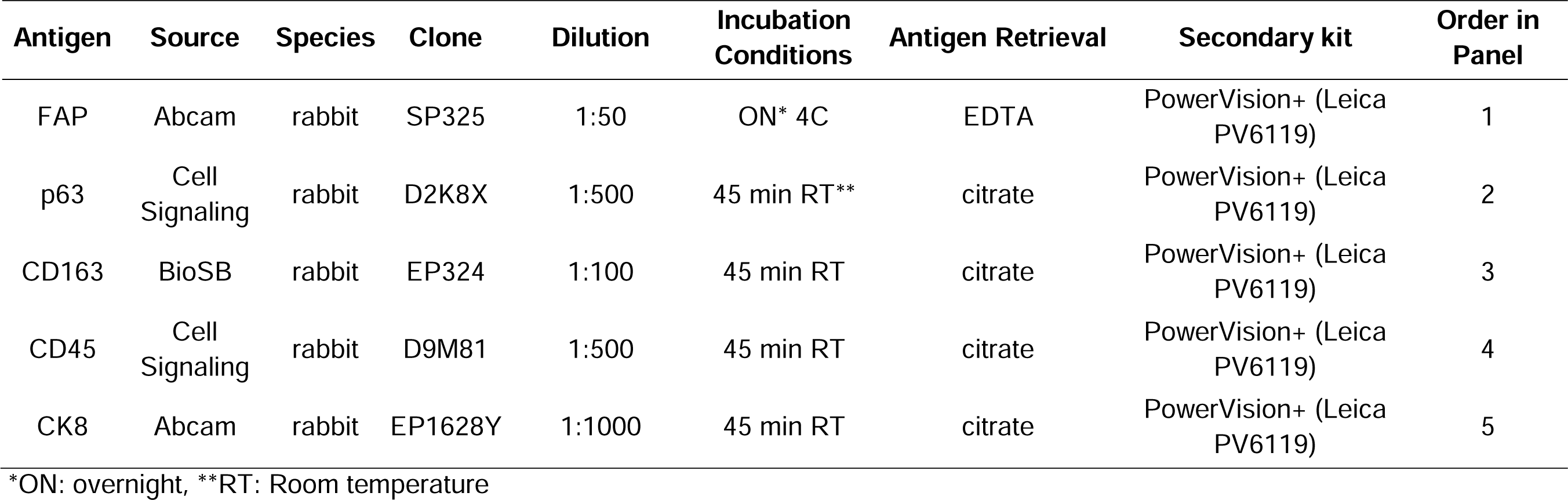
Antibodies, pretreatment and secondary antibody details and staining conditions.

| Antigen | Source | Species | Clone | Dilution | Incubation Conditions | Antigen Retrieval | Secondary kit | Order in Panel |
| --- | --- | --- | --- | --- | --- | --- | --- | --- |
| FAP | Abcam | rabbit | SP325 | 1:50 | ON* 4C | EDTA | PowerVision+ (Leica PV6119) | 1 |
| p63 | Cell Signaling | rabbit | D2K8X | 1:500 | 45 min RT** | citrate | PowerVision+ (Leica PV6119) | 2 |
| CD163 | BioSB | rabbit | EP324 | 1:100 | 45 min RT | citrate | PowerVision+ (Leica PV6119) | 3 |
| CD45 | Cell Signaling | rabbit | D9M81 | 1:500 | 45 min RT | citrate | PowerVision+ (Leica PV6119) | 4 |
| CK8 | Abcam | rabbit | EP1628Y | 1:1000 | 45 min RT | citrate | PowerVision+ (Leica PV6119) | 5 |

\*ON: overnight, \*\*RT: Room temperature

